# Bayesian spatial modelling of Ebola outbreaks in Democratic Republic of Congo through the INLA-SPDE approach

**DOI:** 10.1101/2020.04.13.20063081

**Authors:** Ezra Gayawan, Oyelola A. Adegboye, Adewale James, Adedayo M. Adegboye, Faiz Elfaki

**Author notes:** Correspondence: Oyelola A. Adegboye, Ton Duc Thang University, Ho Chi Minh City, Vietnam. Joint first authors (Ezra Gayawan, Oyelola A. Adegboye).

## Abstract

Ebola virus (EBV) disease is globally acknowledged public health emergence, which is endemic in the West and equatorial Africa. To understand the epidemiology especially the dynamic pattern of EBV disease, we analyse the EBV case notification data for confirmed cases and reported deaths of the ongoing outbreak in Democratic Republic of Congo (DRC) between 2018 and 2019, and examined the impart of reported violence of the spread of the virus. Using fully Bayesian geo-statistical analysis through stochastic partial differential equations (SPDE) that allows us to quantify the spatial patterns at every point of the spatial domain. Parameter estimation based on the integrated nested Laplace approximation (INLA). Our findings reveal strong association between violent events in the affected areas and the reported EBV cases and deaths, and the presence of clusters for both cases and deaths both of which spread to neighbouring locations in similar manners. Findings from the study are therefore useful for hotspot identification, location-specific disease surveillance and intervention.

**Impacts:** In 2018, the Democratic Republic of Congo (DRC) confirmed their tenth Ebola epidemic in 40 years. The outbreak is the country’s largest Ebola outbreak and the second largest ever recorded after the West African epidemic of 2014-2016.

The current outbreak is reported to be occurring in a longstanding conflict zone, this study focused investigating the spatial distribution of Ebola incidence in DRC and the role of violent events.

Violent events in the affected areas was found to be significantly associated with reported Ebola cases, which is highly relevant for hotspot identification and location-specific disease surveillance and intervention.

## 1. Introduction

The ongoing human Ebola virus outbreak in several sub-Saharan African countries is especially unprecedented. Ebola virus (EBV) is a negative-stranded RNA virus of the family Filoviridae that is endemic to regions of the west and equatorial Africa (Malvy, McElroy, de Clerck, Günther, & van Griensven, 2019). EBV is very deadly, with varying fatality rates which could be up to 90% (Dixon & Schafer, 2014; Ilunga Kalenga et al., 2019; Kucharski & Edmunds, 2014; Lefebvre et al., 2014). The 2014 epidemic started when the virus crossed from infected wildlife to human population. Subsequently, it began to spread between people through direct contact with infected blood, secreted bodily fluids, and surfaces of materials contaminated with these secretions (Dixon & Schafer, 2014; Qiu et al., 2013). The risk of transmission is higher when the infected patient is in the later stages of illness because viremia (the presence of the virus in the blood) is higher at these stages (Dowell et al., 1999; Smith, 2009).

The largest outbreak, which challenged the health systems of Sierra Leone, Liberia and Guinea, was recorded between 2014 and 2016 (WHO Ebola Response Team, 2016). More than 28,000 cases and 11,000 confirmed dead, mostly from the populations of these three countries, were recorded (Subissi et al., 2018; WHO Ebola Response Team, 2016). Thus, the general populace, especially the health-care workers in the different West African countries, are exposed to greater risk. A recent study indicated that the short-term (3 and 6 weeks) probability of international spread outside the African continent is small but not negligible and that any further spread in more African countries could enhance the likelihood of international dissemination (Gomes et al., 2014; Pigott et al., 2014).

The current outbreak that begun in August 2018 is mainly concentrated in three provinces located in the North/South Eastern region of the Democratic Republic of Congo (DRC). As of 15 December 2019, the World Health Organization (WHO) had recorded about 3348 EBV cases and 2213 deaths (World Health Organization, 2019b). The outbreak is concentrated in cities and towns housing major domestic and international airports and, it is the tenth and largest in DRC since the first was reported in 1976 (Dyer, 2018; Maxmen, 2018; Wannier et al., 2019).The epidemiology of EBV disease is complex because of the pattern of spread, which is favored by densely populated areas. The rate at which the outbreak is evolving cannot be overemphasized and the geographical extension of its spread is widening which, in part, is due to violence targeted at health workers (Wannier et al., 2019).

There is extensive literature on the relationship between conflict and infectious diseases (O. A. Adegboye & Danelle, 2014; Gayer, Legros, Formenty, & Connolly 2007; Qadri, Islam, & Clemens, 2017; Sharara & Kanj 2014; Stone-Brown, 2013). Armed conflicts provide fertile ground for the emergence and spread of infectious diseases and leads to destruction of resources and health infrastructure, displacement of large parts of the population to crowded refugee camps and thus accelerating infections (O. A. Adegboye & Danelle, 2014). The current EBV outbreak is reported to be occurring in a long-standing conflict zone and it is suspected that there is a relationship between the number of violent attacks and the rate of transmission. The transmission rates between the conflict zones suggest that violent attacks are contributing to the increased transmission of the disease (Wannier et al., 2019). During the World Health Assembly at Geneva, Switzerland, the DRC’s minister for health, Oly Ilunga Kalenga, told reporters that the country’s government is unable to contain the spread of the disease due to the increase in violent attacks against health care facilities. Kalenga stated that “the real emergency we face right now is security” (Bibbo, 2019).

Geostatistical analysis of infectious disease burden is an important approach in disease prevention and control strategies because it provides a graphical explanation of the relationship between disease burden, human population and their geographical place of residence (O. Adegboye, Leung, & Wang, 2018; Manto, 2005). The analysis of the spatial patterns of the current Ebola outbreak would reveal spatial autocorrelation that could rather remain unknown if traditional methods are employed. Several spatial and disease mapping techniques have been proposed and utilized in understanding the spatial distributions of disease burden across different geographical settings (Besag, 1974; Besag, York, & Mollié, 1991; Breslow & Clayton, 1993; Clayton, Spiegelhalter, Dunn, & Pickles, 1998; Cressie, 1992). Models for areal data such as the commonly used intrinsic conditional autoregressive model proposed by Besag (Besag et al., 1991) summarizes disease burden of a geographical entity based on its county or regional division and forces the random effects to exhibit a single global level of spatial smoothness determined by geographical adjacency which is not flexible enough to capture the complex localized structure likely to be present in the residual spatial autocorrelation (Lee, Rushworth, & Sahu, 2014). We map the spatial distribution of the latest outbreak in DRC by adopting a Bayesian kriging approach through the stochastic partial differential equations (SPDE) and parameter estimation was based on the integrated nested Laplace approximation (INLA) proposed by Rue et al. (Rue, Martino, & Chopin, 2009). INLA is becoming popular with complex models in different fields (Schrödle & Held, 2011; Selle, Steinsland, Hickey, & Gorjanc, 2019; Ugarte, Adin, Goicoa, & Militino, 2014) and with SPDE for spatio-temporal geo-statistical data analysis (Abd Naeeim, Abdul Rahman, & Muhammad Fahimi, 2020; Baquero & Machado, 2018; Blangiardo & Cameletti, 2015; Cameletti, Lindgren, Simpson, & Rue, 2013; Mayfield et al., 2018).

For continuous spatial variables, which are, only measured at some finite set of specific points, like the case of the Ebola outbreak, it might be useful to predict their values at some unobserved locations within the geographical unit. This approach might play important role in health risk management in order to understand and identify areas where the risk of exceeding potentially harmful thresholds is higher. Therefore, in the present study, we quantified the extent of the ongoing EBV outbreak in DRC and predict its occurrence at continuous spatial locations. In addition to measuring spatial dependence, we model the effect of violent attacks on the spread of EBV in the affected region. The analysis was performed for reported cases and repeated for recorded deaths. Our findings contributed to expanding the present understanding of the transmission dynamics underlying the outbreak that may be useful in the different frontiers of its upsurge, possibility of its containment and design of control strategies in future.

## 2. Materials and methods

### 2.1 DRC Ebola outbreak and event data

Weekly Ebola epidemic report at specific locations in the health zones (HZ) (Figure S1) of the three affected provinces in DRC between 30 April 2018 and 17 December 2019 were extracted from the publicly available disease outbreak news published by the WHO (World Health Organization, 2019b). The data are elaborate and reliable, reflecting the case notifications from the Ministry of Health, DRC (Wannier et al., 2019). The coordinates of the locations where the cases occurred were obtained and the count data were linked to their point of occurrence. Additionally, we extracted conflict data during the same period from the Armed Conflict Location & Event Data Project (ACLED) (https://www.acleddata.com/) (Raleigh, Linke, Hegre, & Karlsen, 2010), and linked them to the locations where the EBV cases were observed based on proximity. ACLED provide real-time data that capture armed and non-armed conflict events especially in developing countries on Africa and Asia continents. It is also widely used for analysis source on political violence and protest around the world (Raleigh et al., 2010).

### 2.2 Descriptive analysis

We began our analysis with descriptive summaries presented as counts and percentages, and data visualization on maps. We estimated the local indicators of spatial associations (LISA) (Anselin, 1995) using bivariate local Moran’s I statistics to assess the correlation of EBV cases and violent events with reference to spatial location (spatial autocorrelation at each specific health zones). For the spatial weights, we used Queen-style contiguity 1^st^ order nearest neighbour (i.e., two districts are neighbouring if they share common borders or a point).

### 2.3 INLA-SPDE

In the present application, we modelled the observed cases and deaths from EVB at specific locations through the SPDE approach with INLA, controlling for the number of attacks at specific locations. Specifically, we let y_i_ be the reported cases of or deaths from EVB in the study locations. The response variables were considered to have come from Poisson distribution such that

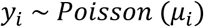

so that

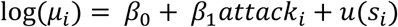

where *β*_0_ is the model intercept, *β*_1_ is the linear parameter for number of attacks and *u*(*s*_*i*_)is the spatial random component for the point-referenced data.

The SPDE approach involves representing the continuously indexed spatial process (Gaussian Field (GF)) with a Matérn covariance function as a discretely indexed spatial random process. Thus, basis function representation provides the link between the GF and Gaussian Markov random field (GMRF) making it easier to use a fast computational numerical methods such as the (INLA) as implemented in the R-INLA package [36].

The Matérn covariance function is given by

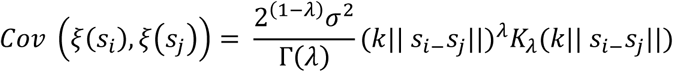

where ∥*s*_*i*−_*s*_*j*_∥ is the Euclidean distance between any two locations *s*_*i*_ and *s*_*j*_, *K*_*λ*_ is a modified Bessel function of the second kind and order *λ*>0 measures the degree of smoothness of the process, *σ*^2^ is the marginal variance, and *k* is a scaling parameter related to the range *r*, which is the distance at which the spatial correlation becomes almost null. The empirical definition is 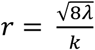

The link between SPDE and the Matérn parameters is given by

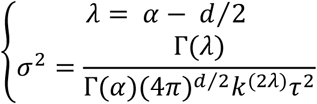

The solution to the SPDE represented by the stationary and isotropic Matérn GF *ξ*(s) are approximated through a basis function representation defined on a triangulation, which divides the spatial domain into a set of non-intersecting triangles, of the domain D:

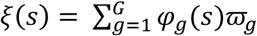

where G is the total number of vertices of the triangulation, *φ*_*g*_ is the set of basis function and 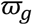 are zero mean Gaussian distributed weights. To ascertain a Markov structure, the basis function are chosen to have a local support and to be piecewise linear in each triangle so that *φ*_*g*_=1 at vertex g and 0 at other vertices (Blangiardo & Cameletti, 2015). In practice, the mesh definition (Figure S2) is a trade-off between the accuracy of the GMRF representation and computational costs that both depend on the number of vertices used in the triangulation.

## 3. Results

### 3.1 Exploratory data analysis

Of the 3351 case notification data from DRC for the period April 2019-December 2019, 3233 were confirmed EBV cases. Table 1 presents the descriptive summaries for the data. During this period, 2099 deaths from EBV were confirmed yielding fatality rate of 64.9%. There were more females (56.3%) infected with EBV than males and about 28.1% of the infected were less than 18 years of age. Figure 1 presents the distribution of EBV cases across health zones in the three affected provinces-North. Note that there was an increasing spread of EBV from the centre of North Kivu northwards. Among the three affected provinces, 84.2% of EBV cases occurred in North Kivu. The burden of the outbreak is mostly concentrated in two health zones in North Kivu: Beni (695) and Katwa (651).

**Table 1.** Descriptive summaries of the EBV notification data, (30 April 2018 to 17 December 2019)

| Characteristics | n | % |
| --- | --- | --- |
| Age group |  |  |
| <18 | 941 | 28.1 |
| 18+ | 2410 | 71.9 |
| Gender |  |  |
| Male | 1465 | 43.7 |
| Female Cases | 1886 | 56.3 |
| Overall | 3351 |  |
| Probable | 118 | 3.5 |
| Confirmed | 3233 | 96.5 |
| North Kivu | 2723 | 84.2 |
| Ituri | 504 | 15.6 |
| South Kivu | 6 | 0.2 |
| Deaths | n | %CFR |
| Overall | 2217 | 66.1 |
| Probable | 118 | 100.0 |
| Confirmed | 2099 | 64.9 |
| North Kivu | 1847 | 67.8 |
| Ituri | 249 | 49.4 |
| South Kivu | 3 | 50.0 |

**Figure 1.**
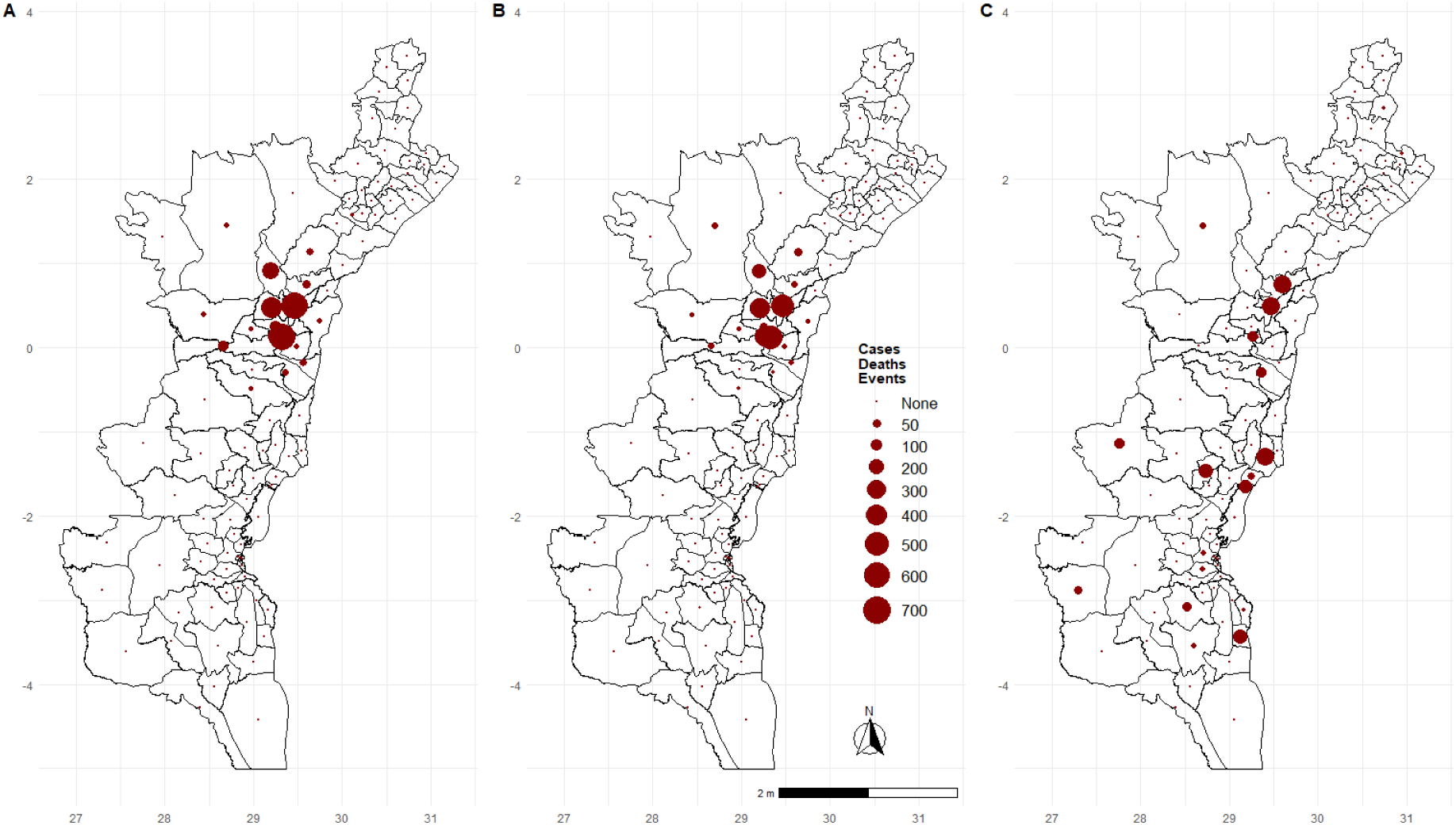
Map showing number of (A) Ebola cases, (B) Deaths, (C) Violent events, across Health zones in DRC between April 2018 and December 2019.

Figure 2 presents the LISA maps of the EBV cases and recorded violent events in the study area. It is evident from the LISA maps, that there exist spatial associations between number of violent events and cases of (and deaths from) EBV diseases in DRC. We observed significant clustering of high EBV deaths and high violent events in Oicha but low EBV deaths and high violent events in HZ in north-eastern parts of North-Kivu. We found significant clusters of high EBV cases and high violent events in Mambasa and Oicha (Figure 2), both in Ituri and North-Kivu provinces, respectively. Considerable number of clusters of low EBV cases and high violent events was observed in North-Kivu.

**Figure 2.**
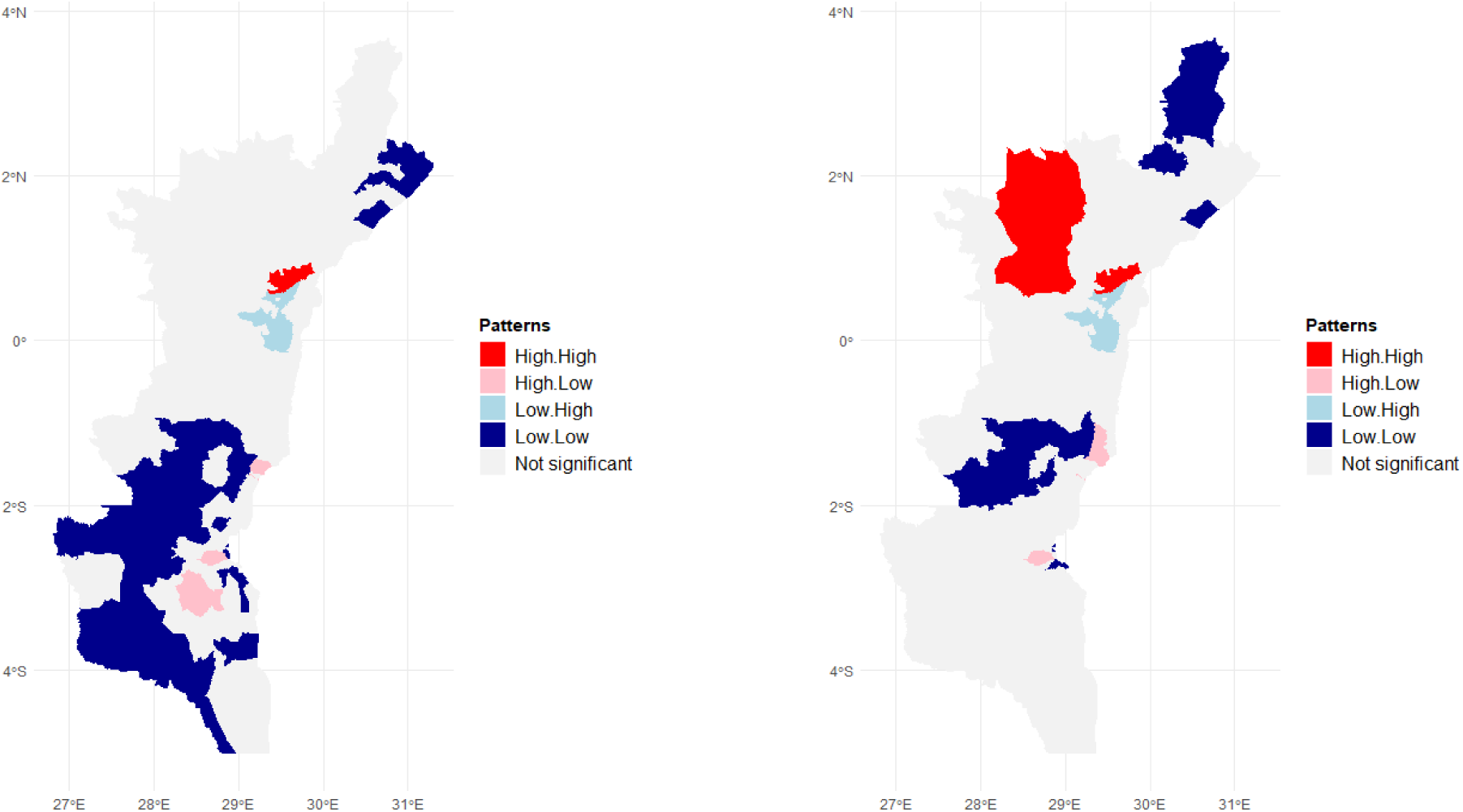
LISA map of bivariate Moran’s I spatial correlation. Mortality. Right: Cases vs. events, Left: Mortality vs. events.

### 3.2 Hierarchical models

We consider four hierarchical models and made comparison based on Deviance Information Criterion (DIC) where the model with least DIC value is considered the best. The first was a null model containing no covariate. The second includes the violent event as a fixed linear effect while the third has only the spatial random effect. The last model considers both the linear and spatial effects. Table 2 presents the values of the model diagnostic criterion showing the mean deviance 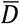, effective number of parameters (pD), which is similar but not equal to the degree of freedom and DIC values. For both confirmed EBV cases and deaths, the inclusion of spatial effect improved the fit when compared with the other two models. However, the difference in DIC values for the third and fourth model is less than 10 and so, none can be adjourned to be better than the other, and normally, the model with fewer parameters should be embraced in such situation. In order to evaluate the effects of the conflict variable, we present results of the last model as contained in Table 3 and Figure 3.

**Table 2.** Comparison of model’s statistics.

| Model | $D^-$ | pD | DIC |
| --- | --- | --- | --- |
| Deaths |  |  |  |
| Null | 10522.96 | 1.01 | 10551.95 |
| Fixed effect, no spatial | 9230.92 | 2.01 | 9228.81 |
| Spatial only | 271 | 71.03 | 199.97 |
| Fixed effect + spatial | 263.64 | 67.31 | 196.34 |
| Cases |  |  |  |
| Null | 14901.94 | 1.02 | 14900.92 |
| Fixed effect, no spatial | 13075.08 | 2.02 | 13073.06 |
| Spatial only | 308.43 | 79.43 | 229 |
| Fixed effect + spatial | 299.85 | 75.09 | 224.75 |

**Table 3.** Posterior distribution (mean, sd and quantiles) of the model parameters of the INLA-SPDE models.

| Parameter | Mean | SD | 2.5% percentile | 97.5% percentile |
| --- | --- | --- | --- | --- |
| Deaths |  |  |  |  |
| Intercept, $\alpha$ | -4.448 | 1.147 | -7.060 | -2.577 |
| Violent event, $\beta$ | 0.022 | 0.009 | 0.005 | 0.041 |
| $\tau$ | 0.004 | 0.002 | 0.003 | 0.005 |
| Scale parameter, $\kappa$ | 17.329 | 7.481 | 13.873 | 22.446 |
| Variance, $\sigma^2$ | 18.444 | 5.926 | 15.243 | 22.606 |
| Practical range, $r$ | 0.163 | 0.057 | 0.125 | 0.203 |
| $\vartheta_1$ | -5.611 | 0.394 | -6.464 | -4.931 |
| $\vartheta_2$ | 2.855 | 0.355 | 2.268 | 3.652 |
| Cases |  |  |  |  |
| Intercept, $\alpha$ | -4.585 | 1.223 | -7.378 | -2.581 |
| Violent event, $\beta$ | 0.024 | 0.010 | 0.005 | 0.045 |
| $\tau$ | 0.003 | 0.001 | 0.002 | 0.004 |
| Scale parameter, $\kappa$ | 18.523 | 7.919 | 14.726 | 23.955 |
| Variance, $\sigma^2$ | 24.230 | 7.238 | 19.950 | 29.320 |
| Practical range, $r$ | 0.152 | 0.055 | 0.117 | 0.192 |
| $\vartheta_1$ | -5.611 | 0.399 | -6.666 | -5.106 |
| $\vartheta_2$ | 2.944 | 0.360 | 2.293 | 3.702 |

**Figure 3.**
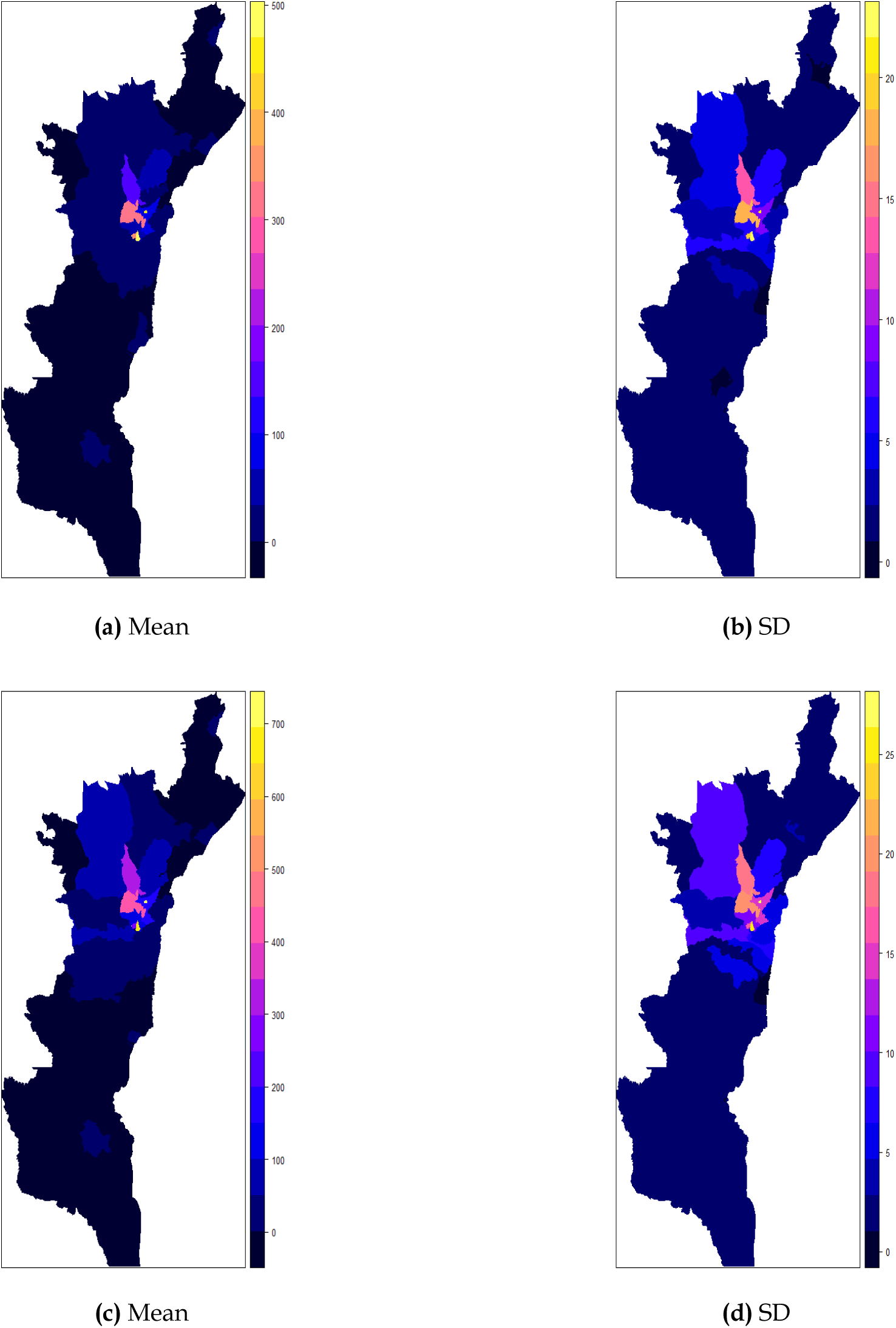
Spatial distribution of posterior means and standard deviations using INLA-SPDE (top left and top right, respectively). (Top) Deaths, (Bottom) Cases.

The marginal distributions of the parameters of the INLA-SPDE models including the scale parameter (*κ*) of the Matérn function, variance parameter of the random field (*σ*^2^) and the practical range, (*r*) are presented in Table3. For example, to predict confirmed EBV cases, the posterior mean of Matérn function are: scale parameter (15.523), variance (24.230) and range (0.152). The marginal distributions of the fixed effect (number of violent events) indicates a positive significant association with EBV cases and deaths. The posterior distributions for fixed and hyper-parameters for both models are displayed in Figures S3-S6.

Figure 3 shows the spatial distributions of the posterior mean and standard deviation (uncertainty due to the model parameter) of predicted EBV cases and deaths. The maps show that predicted EBV deaths and cases are higher and concentrated in the bordering health zones between Ituri and North Kivu, respectively after accounting for the number of violent events (Figures 3a & 3c). The spatial variations of the standard deviations are similar to the posterior mean for both cases and deaths. They range between 0 to 25 for EBV deaths and 0 to 30 for EBV cases. The uncertainty spread out from the same bordering health zones (Figures 3b & 3d).

## 4. Discussion

It is known that conflicts and social instability are among the major risk factors for emerging and re-emerging infectious diseases (Nii-Trebi, 2017). Provinces in DRC have been plagued by decades of violence by rebel groups and the recent outbreak of Ebola in the North/South Eastern areas of the country has worsen the situation. Absence of adequate security provide fertile ground for the re-emergence and spread of infectious diseases such as Ebola. This study used Bayesian SPDE approach to generate continuous spatial maps of EBV cases and deaths while accounting for the influence of violent events in DRC. Our results confirmed that the current outbreak of Ebola disease varied spatially with higher concentrations at certain locations and that spreads to neighbouring locations. Also, as expected, the pattern of the spatial spread of confirmed EBV cases is similar to that of recorded deaths. Findings from a recent study (Richardson et al., 2016), suggest that the role of human rights failings such as colonial legacies, structural adjustment, exploitative mining companies, enabled civil war, rural poverty, and the near absence of quality health care could account for the genesis of the 2013-2016 Ebola outbreak and same could have been applicable to the current outbreak. Further, high level of poverty in the country leaves the populace to expand their reach for food and livelihood by encroaching deeper into the forests thereby, enhancing their risk of exposure to Ebola virus and other zoonotic pathogens (Bausch & Schwarz, 2014). All these take place in the face of a weak and dysfunctional health care system (Tomori, 2014).

We found a positive significant association between number of violent events and EBV cases and deaths. This finding is especially intuitive for Ebola disease since violence leads to destruction of health resources and infrastructure, as well as displacement of population which potentially could exacerbate infections. In the last week of November 2019, several Ebola response operations were paralysed due to attacks on the facilities by armed group. These actions hinder adequate and timely response by the EBV response personnel who could administer the needed vaccination. The natural outcome of such action is potential increase in the transmission and rise in the number of cases due to impacts on surveillance and control efforts such as active case finding, contact tracing, and vaccination (World Health Organization, 2019a). The number of internally displaced persons (IDPs) in the EBV affected areas is spiralling; more than 100,000 in North-Kivu, 227,000 in Ituri and 263,252 IDPs in South Kivu (as at 22 December 2019) (World Health Organization, 2019c). These huge concentration of persons in IDP camps lead to closer contacts between humans and this exacerbates the risks of the infection.

We identified the following limitations in this study. We did not distinguished the type of violent event as impacting directly or indirectly the health-care systems in the affected areas. We captured the event-EBV relationship as nonlinear, there is possibility that effect violent events may not be immediate, therefore, a spatial lag nonlinear time series model may be desirable. Lastly, other mitigating factors such environmental factors and socioeconomic factors could be considered in future studies.

## 5. Conclusion

The use of the SPDE approach based on continuous spatial random field to study the spatial distributions of the EBV outbreak in parts of DRC has allowed us to quantify the spread at every point of the spatial domain and thus allowing the proper view of the impact at every location including places where data were not collected. This approach is more advantageous than adopting a method that sums the disease occurrence based on geographical areas. We have expanded on the dynamics of the ongoing EBV outbreak in DRC by predicting its occurrence at continuous spatial locations and in particular, the effects of the conflicts in the region. Findings from the study are therefore useful for hotspot identification, location-specific disease surveillance and intervention, control strategies, health planning and funding allocation for curtailing the disease. Proper disease surveillance and response systems need to be strengthened in the affected areas and indeed in all parts of the country to enhance early detection and control thus, curtailing further spread whether now or in the future.

## Data Availability

Publicly available disease outbreak news published by the WHO

## Appendices

**Figure S.1.**
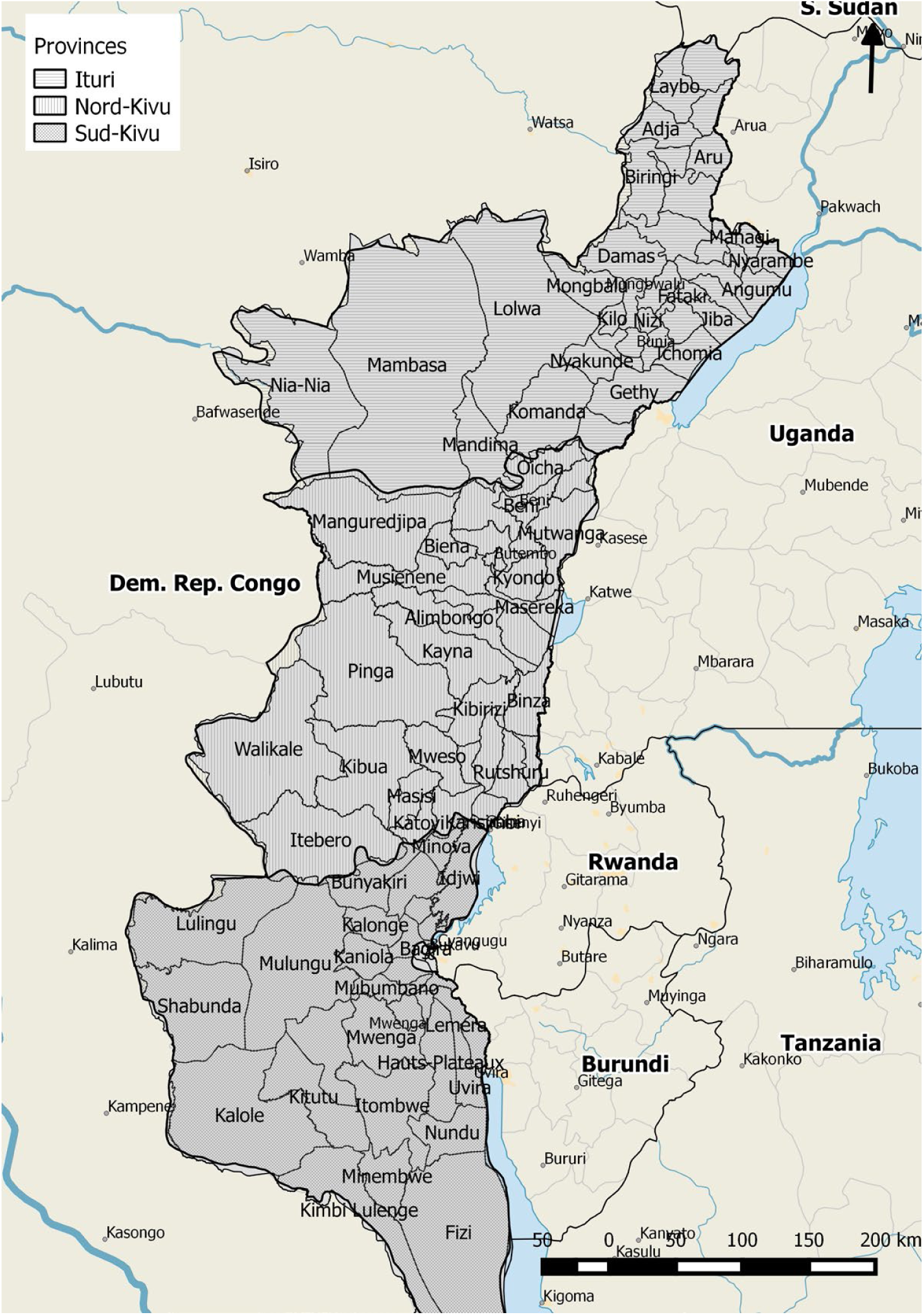
Map of DRC showing the outbreak provinces and health zones.

**Figure S.2.**
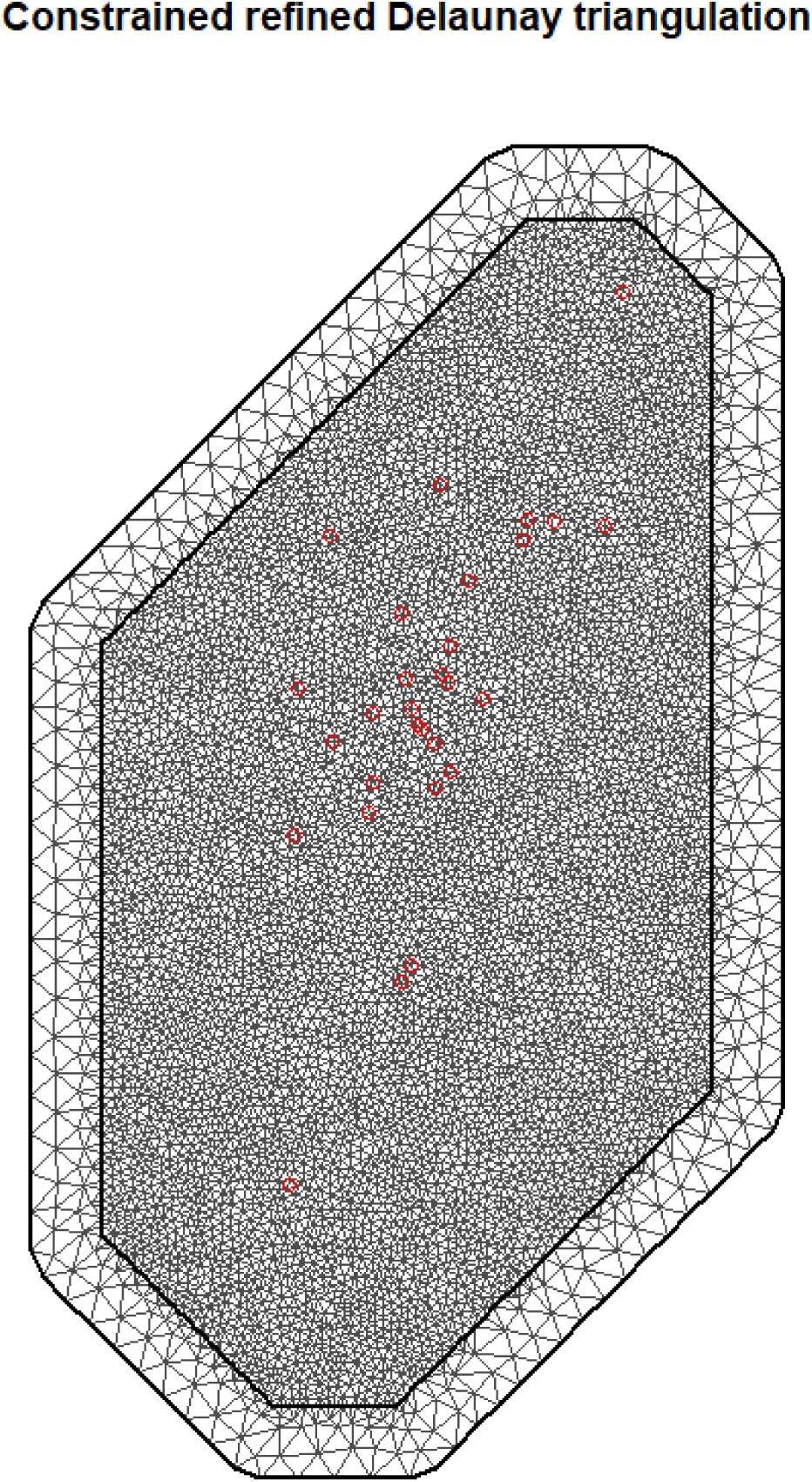
Delaunay triangulation (mesh) of Ebola incidence (red dots) in the study area.

**Figure S.3.**
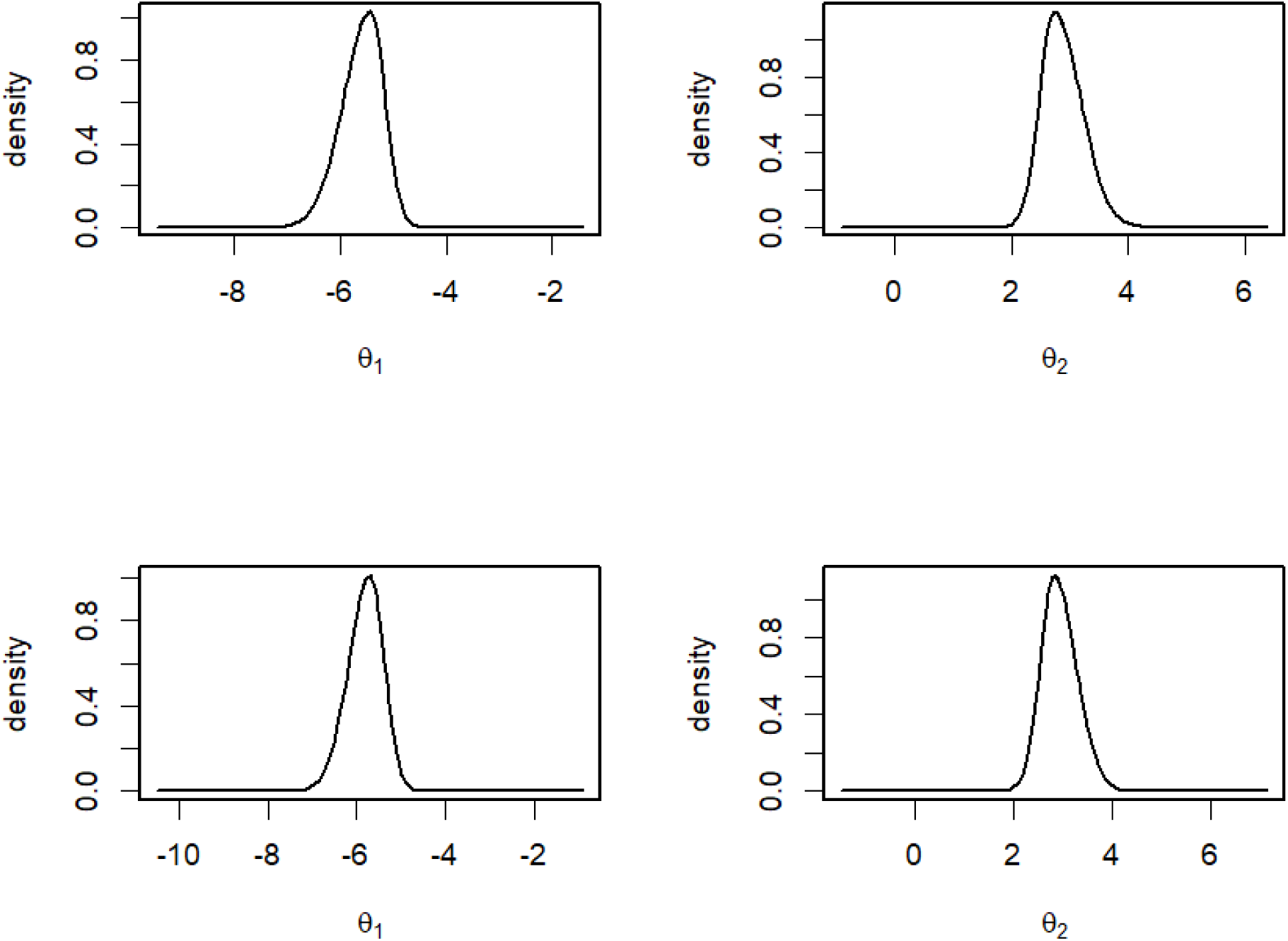
Posterior marginal distribution for *theta*_1_ and *theta*_2_. (Top) Deaths, (Bottom) Cases.

**Figure S.4.**
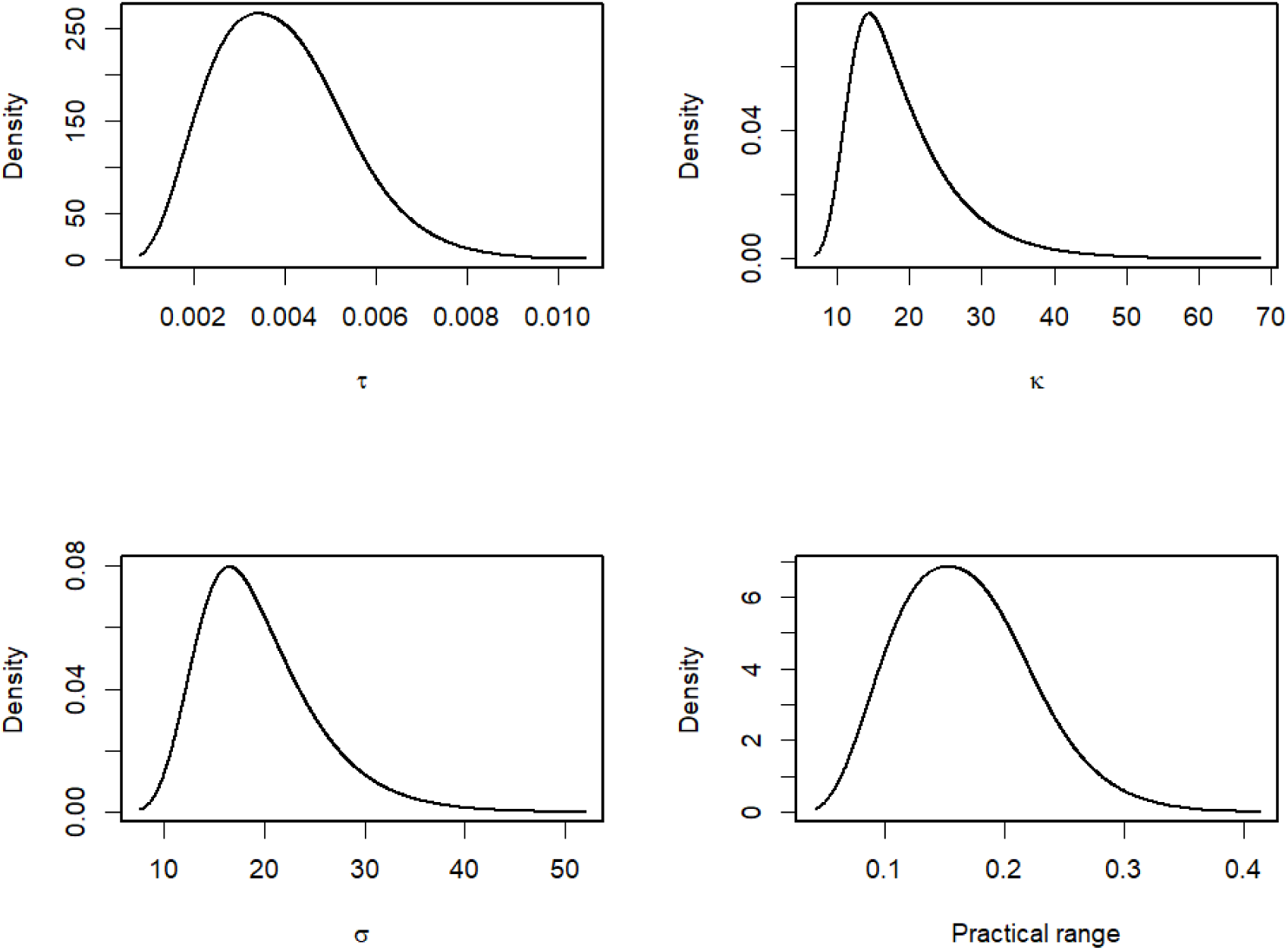
Posterior marginal distribution for hyper-parameters in the mortality (deaths) model.

**Figure S.5.**
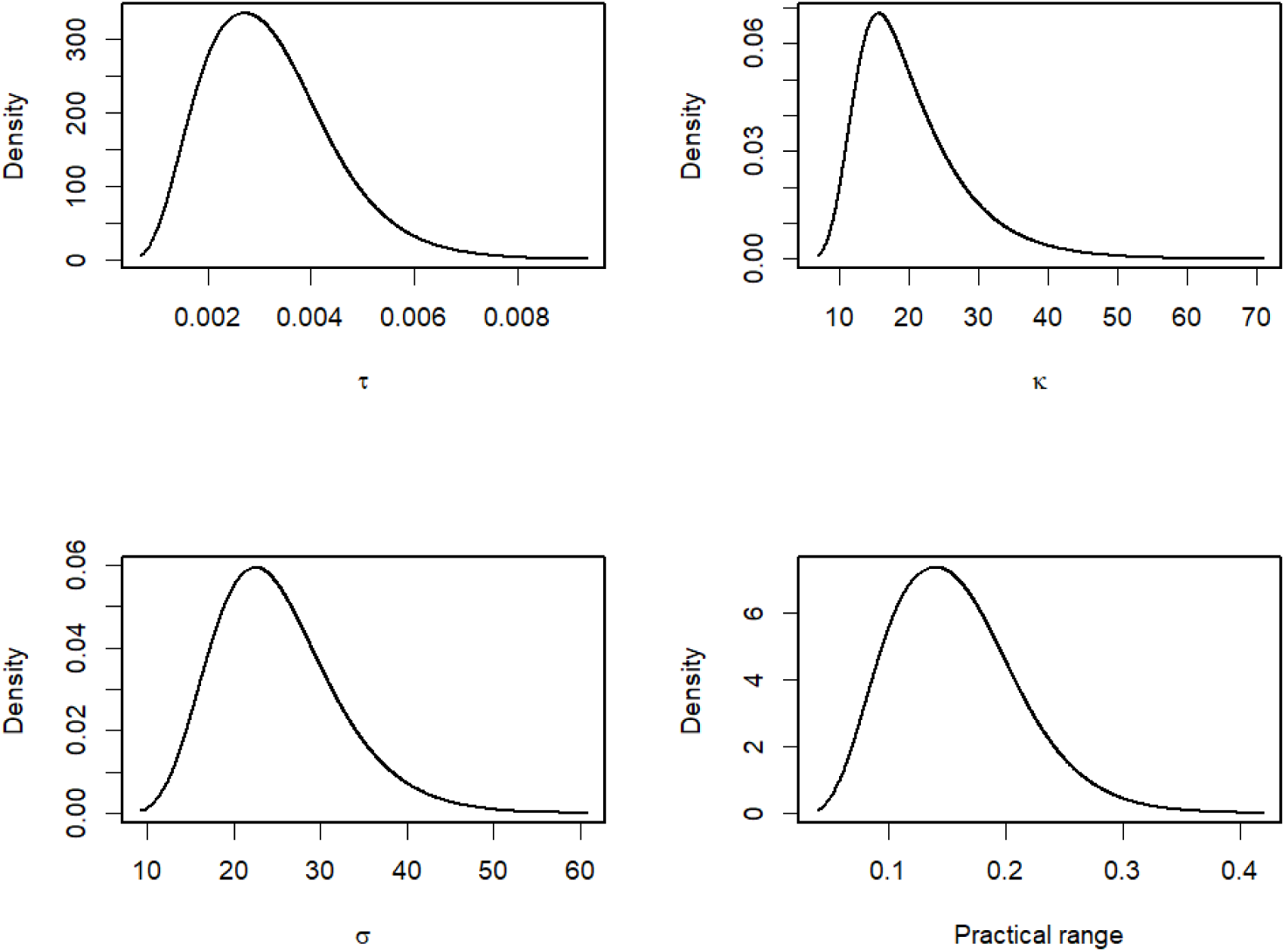
Posterior marginal distribution for hyper-parameters in the incidence (cases) model.

**Figure S.6.**
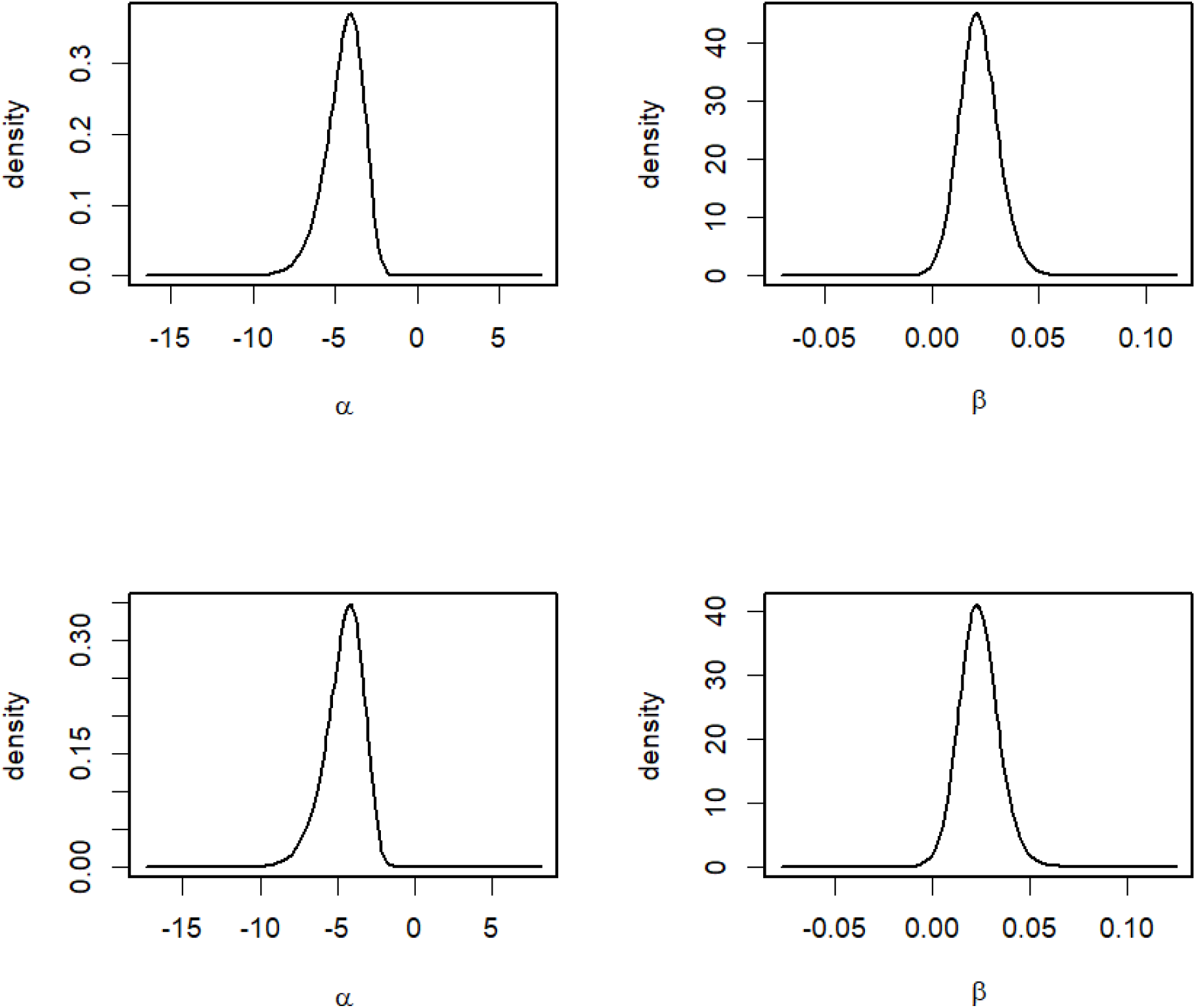
Posterior distribution of the intercept and the covariate coefficient. (Top) Deaths, (Bottom) Cases.

